# Repurposing Existing Medications for Coronavirus Disease 2019: Protocol for a Rapid and Living Systematic Review

**DOI:** 10.1101/2020.05.21.20109074

**Authors:** Benjamin P. Geisler, Lara Zahabi, Adam E. Lang, Naomi Eastwood, Elaine Tennant, Ljiljana Lukic, Elad Sharon, Hai-Hua Chuang, Chang-Berm Kang, Knakita Clayton-Johnson, Ahmed Aljaberi, Haining Yu, Chinh Bui, Tuan Le Mau, Wen-Cheng Li, Debbie Teodorescu, Ludwig Christian Hinske, Dennis L. Sun, Farrin A. Manian, Adam G. Dunn

## Abstract

**Background:** Coronavirus Disease 2019 (COVID-19) has no known specific treatments. However, there might be *in vitro* and early clinical data as well as evidence from Severe Acute Respiratory Syndrome and Middle Eastern Respiratory Syndrome that could inform clinicians and researchers. This systematic review aims to create priorities for future research of drugs repurposed for COVID-19.

**Methods:** This systematic review will include *in vitro*, animal, and clinical studies evaluating the efficacy of a list of 34 specific compounds and four groups of drugs identified in a previous scoping review. Studies will be identified both from traditional literature databases and pre-print servers. Outcomes assessed will include time to clinical improvement, time to viral clearance, mortality, length of hospital stay, and proportions transferred to the intensive care unit and intubated, respectively. We will use the GRADE methodology to assess the quality of the evidence.

**Discussion:** The challenge posed by COVID-19 requires not just a rapid review of drugs that can be repurposed but also a sustained effort to integrate new evidence into a living systematic review.

**Systematic review registration:** PROSPERO 2020 CRD42020175648

## Background

The Coronavirus Disease 2019 (COVID-19) pandemic represents one of the deadliest and economically most consequential outbreaks in around 100 years [1, 2]. To date, only remdesivir (Gilead Sciences) has shown to possibly lower the time to recovery [3, 4]. In the past, developing antiviral agents has taken 5.9 years on average [5]. In the current crisis, it appears feasible that this timeline will be significantly reduced if there were early signs of efficacy. Moreover, a candidate drug could be offered emergency use authorization by the U.S. Food and Drug Administration and similar designations by regulatory authorities across the world, as was the case with remdesivir.

Drug repurposing has been a successful strategy for a variety of therapeutic areas [6]. In the case of COVID-19, compounds of interest include those that have previously been found to either be clinically efficacious or have *in vitro* activity against the coronaviruses that cause Severe Acute Respiratory Syndrome (SARS) and Middle Eastern Respiratory Syndrome (MERS), which share significant structural similarities with SARS-CoV-2, the virus that causes COVID-19 [7, 8]. Information on drugs potentially active against COVID-19 is expected to change rapidly as the results of ongoing and future studies become available. It is crucial that both researchers and health care providers are able to access the optimum and most up-to-date information to inform future or ongoing studies and provide clinical care.

The objectives for the current protocol are (1) to systematically review which existing medications have shown to be potentially effective against SARS or MERS and could therefore be potentially repurposed; (2) to present the early evidence that supports testing readily available drugs against SARS-CoV-2; (3) to provide the best available level of evidence for the efficacy of each individual candidate drug; and (4) to report harmful effects associated with use of these drugs.

## Methods/Design

This protocol specifies the conduct and reporting of a systematic review in compliance with the Preferred Reporting Items for Systematic Review and Meta-Analysis Protocols 2015 statement (PRISMA-P) [9]. We intended to complete the review within 60 to 90 days. The protocol has been registered with the International Prospective Register of Systematic Reviews (PROSPERO) and assigned the identifier CRD42020175648. Beyond a traditional systematic review, we aim to make this a living document (see “Living Systematic Review” below).

### Data sources

Bibliographical databases for literature search include Medline (via the Entrez PubMed interface), Embase (via the Embase.com interface), ClinicalTrials.gov (including studies that have already posted results), and Google Scholar. We also target the following preprint servers: MedRxiv, BioRxiv, chemRxiv, Preprints.org, and the Chinese-language server ChinaXiv. Our search strategy combines terms for COVID-19, SARS, MERS, and their causative agents with drug names, based on the following eligibility criteria (see also Appendix).

### Eligibility Criteria

The retrieved studies will be selected according to the eligibility criteria listed below.

#### Study Design

*In vitro* studies, animal studies, and clinical studies are all eligible study designs. Single-arm and controlled studies with or without randomization as well as open-label or blinded designs will be included.

#### Interventions

The interventions we studied were derived from a scoping review by the World Health Organisation (WHO) [10]. They encompass both single-agent and combination regimens with one or more of the following agents: atazanavir, azithromycin, baloxavir marboxil, baricitinib, bevacizumab, chloroquine, colchicine, darunavir/cobicistat, emtricitabine/tenofovir, enisamium iodide, favipiravir (T-705), fingolimod, ganciclovir, hydroxychloroquine, indinavir, lopinavir/ritonavir, mycophenolic acid/mofetil, nelfinavir, niclosamide, nitazoxanide, nitric oxide, novaferon, oseltamivir, pirfenidone, quercetin, remdesivir (GS-5734), ribavirin, ruxolitinib, sirolimus, sofosbuvir, tocilizumab, thymosin alpha-1, triazavirin, and umifenovir. We were also interested in drugs belonging to the following groups: glucocorticosteroids (with or without mineralocorticoids), interferons, and statins. Lastly, we targeted studies on convalescent serum or plasma.

#### Comparators, Participants, and Follow-up Periods

Clinical studies that we seek to evaluate can be either a single-arm study or controlled study, and include patients with mild-to-moderate or severe disease in both outpatient and inpatient settings, with the latter including intensive care unit (ICU) and non-ICU patients. For clinical studies to be included, participants must not just have symptoms compatible with COVID-19 but either laboratory confirmation of infection of respective viruses – Severe Acute Respiratory Syndrome Coronavirus 2 (SARS-CoV-2), Severe Acute Respiratory Syndrome Coronavirus (SARS-CoV), or Middle Eastern Respiratory Syndrome Coronavirus (MERS-CoV) – or be presumed to be infected on clinical grounds after careful deliberation. We do not place any restrictions on minimum follow-up periods.

### Outcomes

We consider the most important study endpoints to include the following variables: time to clinical improvement, time to viral clearance, mortality, hospital admission or transfer to a higher level of care, including transfer to an ICU or ICU-like setting and total and ICU length of stay, proportion of intubations, length of mechanical ventilation, normalization of selected laboratory data (such as C-reactive protein, d-dimer, lactate dehydrogenase, ferritin, and lymphocyte count), and adverse events.

### Study Selection

Two reviewers will use the same eligibility criteria to evaluate the studies. Conflicts will be resolved by discussion.

### Data Extraction

Data from the studies selected for inclusion will be extracted by one reviewer and verified by another. Our team includes reviewers that can read scientific papers a variety of languages, including Chinese. In addition to the outcome measures, the following characteristics of the verified RCTs will be extracted: (1) reference details (including the first author’s last name and the publication year); (2) country of origin; (3) the specific coronavirus studied (SARS-CoV-2, SARS-CoV, or MERS-CoV); (4) type of cell or animal studied (applicable only to *in vitro* or animal study); (5) blinded versus open-label design (applicable to controlled studies); (6) inclusion and exclusion criteria; (7) baseline patient characteristics, in particular duration since onset of symptoms, outpatient versus non-ICU versus ICU setting, and mild-to-moderate versus severe disease; (8) intervention(s) studied; (9) control(s, if any); (10) total number of participants; (11) primary outcome; (12) secondary outcome(s); (13) results; and (14) conclusions.

### Quality assessment

We will assess the design, execution, and reporting of the included studies based on the approach suggested by the Grading of Recommendations Assessment, Development and Evaluation (GRADE) Working Group [11].

### Data synthesis and analysis

The extracted data will be collated and qualitatively synthesized via an interactive mechanism to select, sort, and filter columns for the results tables on our website, www.CovidDrugs.org. If more than one controlled study is available for a similar patient population, we will calculate a relative risk with a 95% confidence interval via meta-analysis in RevMan or STATA. Sensitivity analysis based on the GRADE score will be undertaken if there is evidence of both high and low quality.

### Living Systematic Review

Given that COVID-19 will remain a substantial problem for the foreseeable future, there will be an ongoing need to identify, review and critically appraise [12], and synthesise new studies. We aim to transform this project into a living systematic review [13-16]. We will combine all inputs from the literature databases and the preprint servers into one extensible markup language (XML) stream that we will be made publicly available. As more controlled studies become available, we aim to automate meta-analyses and inform stakeholders once the direction for effect of one of the studied drugs changes (see below). Finally, we will adapt and use methods from natural language processing and crowd-sourcing to support the review process, learning from recently developed tools and processes [17-19]. We will make our methods and results open source and intend to crowd-source future review efforts, which may alter the number of authors on review updates. For permanency, history will be kept and made permanently available via our website www.CovidDrugs.org. The completed review as well as intermittent updates (e.g., when the number of included studies changes, when a conclusion for any intervention changes, or upon some other well-defined trigger) will also be available via the pre-print server medRxiv.

## Discussion

In summary, this systematic review will summarize the emerging evidence on repurposed drugs for the treatment of COVID-19, extrapolating from early evidence for COVID-19 itself as well as SARS and MERS. The protocol represents a rather generic approach to studying the potential treatment compounds identified in previous scoping review by the WHO, except we will search pre-print servers. The initial synthesis will occur rapidly and be more likely than not be qualitative, at least while the evidence is sparse. The present protocol was written in accordance with the PRISMA-P statement. It is registered with PROSPERO. The quality of the evidence will be assessed using the GRADE approach.

The challenge posed by COVID-19, however, requires not just a rapid review of drugs that can be repurposed but also a sustained effort to integrate new evidence into a living systematic review. The outlined approach should be particularly useful in prioritizing drug candidates. Finally, future investigators could benefit from preliminary estimates of magnitudes of the effect of efficacious drugs for power and sample size calculations where appropriate, and their study designs could be informed in terms of endpoints studied and adverse events monitored.

## Data Availability

For permanency, history will be kept and made permanently available via our website http://www.CovidDrugs.org. The completed review as well as intermittent updates (e.g., when the number of included studies changes, when a conclusion for any intervention changes, or upon some other well-defined trigger) will also be available via the pre-print server medRxiv.

**List of Abbreviations**
COVID-19: Coronavirus Disease 2019
GRADE: Grading of Recommendations Assessment, Development and Evaluation
MERS: Middle Eastern Respiratory Syndrome
MERS-CoV: Middle Eastern Respiratory Syndrome Coronavirus
PRISMA-P: Preferred Reporting Items for Systematic Review and Meta-Analysis Protocols
PROSPERO: International Prospective Register of Systematic Reviews
SARS: Severe Acute Respiratory Syndrome
SARS-CoV: Severe Acute Respiratory Syndrome Coronavirus
SARS-CoV-2: Severe Acute Respiratory Syndrome Coronavirus 2
XML: Extensible Markup Language
WHO: World Health Organisation

## Declarations

### Ethics approval and consent to participate

Not applicable

### Consent for publication

Not applicable

### Availability of data and materials

All data and materials will be available at www.CovidDrugs.org.

### Authors’ Contributions

BPG conceived the study, developed the criteria, searched the literature, and wrote the protocol. LZ, AEL, NE, ET, LL, ES, HHC, CBK, KCJ, AA, HY, WCL, and FM assisted in protocol design, managing the literature, selecting the studies, and extracting data. BPG, CB, TLM, DT, LH, DLS, and AGD developed the concept for the automation systems and implemented the website. AGD advised on protocol design and revised the manuscript. All authors read and approved the final manuscript. The opinions or assertions contained herein are the private views of the co-authors and are not to be construed as official or as reflecting true views of their employers.

## Acknowledgements

The authors would like to thank Lisa Liang Philpotts, MSLS of Massachusetts General Hospital, Boston, MA, USA for reviewing an early version of the search strategy.

